# Maternity waiting home Utilization and associated factors among women who gave birth in the Digelu and Tijo district of the Arsi Zone, Oromia, Ethiopia

**DOI:** 10.1101/2020.12.27.20248893

**Authors:** Derese Teshome, Muluemebet Abera, Mamo Nigatu

## Abstract

**Background:** Maternity Waiting Homes (MWHs) is an intervention designed to reduce maternal and perinatal mortality. Ethiopia has introduced the intervention before three decades however; its utilization is very low. Therefore, this study is aimed to assess MWH utilization and associated factors among women who gave birth in the last 12 months in Digelu and Tijo district Arsi Zone Oromia Region, Ethiopia.

**Methods:** Community-based cross-sectional study was conducted in April 2019 on 530 randomly selected women. Data were collected by face-to-face interview using structured questionnaire. Descriptive statistics and logistics regressions were used to analyze the results. Adjusted odds ratio and 95% confidence interval were respectively calculated to measure strength of association and its statistical significance.The confidence interval was used to declare statistical significance in the final model.

**Results:** One hundred twenty-five (23.6%) of the respondents used maternity waiting home. Traveling time less than and equals to 60 minutes from a nearby health facility (AOR=0.16, 95% CI: 0.09, 0.27), women’s decision power (AOR=1.81, 95% CI: 1.10, 2.96), not utilizing antenatal care (AOR=0.6, 95% CI: 0.37, 0.97) and delivering more than three children (AOR=0.56, 95% CI: 0.34, 0.90) were independently associated with utilizing the maternity waiting home.

**Conclusion:** Even though the MWH was designed to reduce maternal and perinatal mortality, less than a quarter (23.6%) of women delivered in the last 12 months before the study in the Digelu and Tijo District had utilized the services. Increasing availability of the service, promoting antenatal care utilization, empowering women and evolving policy makers are recommended to enhance the current low utilization of the MWH.

## Background of the study

Maternity waiting homes (MWHs) have been existed since the early 20th century in the world, particularly in remote rural areas where women have limited access to an obstetric facility. It introduced into developing countries in the 1960’s. MWHs are residential facilities located near a hospital or a health center to accommodate women in their final weeks of pregnancy to bridge the geographical gap (1–3). According to World Health Statistics 2015 released by the World Health Organization every year some 303,000 women die of complications during pregnancy or childbirth globally. About 830 women die from pregnancy- or childbirth-related complications around the world every day. Developing regions accounted for approximately 99% (302 000) of the estimated global maternal deaths in 2015, and Sub-Saharan Africa alone roughly accounted for 66% (201 000) (4). The establishment of MWH has contributed to reduction of maternal mortality ratio and the stillbirth rate(5–7). According to the 2016 Ethiopian Demographic and Health Survey (EDHS), only 28% of delivering women had attended skilled birth attendants(SBA) at national level whereas, only one in five (20%) delivering women has attended the SBA in the Oromia region; Although the maternal mortality ratio has decreased in Ethiopia, it is still very high and access to the emergency obstetrical care is still limited(4). From the study done at Jinka Hospital, southern Ethiopia, 16.7% of the total 516 mothers who were delivered at Jinka hospital were admitted to the MWHs. The MWH was established in 2006; however, it was not well-utilized. Almost all the mothers from the MWHs (98.8%) had delivered at the hospital. Women who had used the MWHs were less likely to experience prolonged PROM, intrauterine fetal death (IUFD) or stillbirth, obstructed labor, Uterine rupture and maternal death(2,8,9). The Maternity waiting home (MWH) utilization has reduced the maternal risk of perinatal death and other related social and economic impact by nearly 50 % as compared to the risk among those who did not utilized it. If women manage to stay in a MWH, immediate access to emergency obstetric care, if required, should be guaranteed(7,10). Studies done at the different countries and few studies from Ethiopia revealed that the utilization of the MWH utilization decreases the maternal morbidity and mortalities(7,11–13). Even though the MWH utilization is proven effective in reducing the maternal and perinatal mortality by different studies done internationally and in Ethiopia, as to the best knowledge of the investigators, its utilization and associated factors are not well studied in the Digelu and Tijo district of Arsi Zone, Ethiopia. Therefore, the current study is aimed at determining the magnitude of the MWH utilization and its associated factors among women who gave birth within the last 12 months at Digelu and Tijo district, South East of Ethiopia, which may be used as baseline for stakeholder to solve the problem.

## Methods and Materials

### Study setting and period

A Community-based cross-sectional study was conducted on women who gave birth in the last 12 months in Digelu and Tijo district. The Digelu and Tijo district is one of the districts in the Arsi Zone of Oromia Region, Ethiopia. It is located at 183 km from Addis Ababa to the south-east. Administratively; the district was subdivided into 28 rural and 2 urban Kebeles which is the lower administrative unit in Ethiopia. The total population of the District, as projected from the 2007 census, was 190,814 of which 88.4% were rural residents, 50.9% were females and 22.9% were women of childbearing age. The district has 5 governmental health centers while 4 of which health centers have the MWHs, while two of them were functional. The district has also currently 23 functional health posts, 49 rural health extension workers, 42 Diploma nurses, 13 midwifes, 7 BSc nurses and 10 health officers. The study was conducted in April, 2019.

#### Inclusion criteria

Women who gave birth within the last 12 months and live more than 6 months in the Digalu and Tijo district.

#### Exclusion criteria

Those women who had difficulty in communication due-to severe illness or unable to respond.

#### Sample size determination and Sampling procedure

The sample size was determined using epi-info version 7.2.2.6, with the assumptions of 95% confidence level, 5% margin of error, 80% power, a design effect of 1.5 and after reviewing different similar studies, 38.7% proportion of MWHs utilization from the study done at Jimma (8) selected which yields highest sample size. Then, the Sample size was calculated to be 507, and by considering a 10% of non-response rate, the final sample size was 558 women. There are 28 kebeles in the district (23 rural and 5urban), and all the kebeles (the lowest administrative level) were stratified into urban and rural kebeles. A total of 9 kebeles, 2 from the urban kebeles and 7 from the rural kebeles were randomly selected. Based on the reports from the HEWs and kebele administrators, there were a total of 1832 households with eligible women in the selected 9 kebeles. The study participants were selected by systematic random sampling with a sampling interval of three households. The first household was randomly chosen by drawing a random number. In a case selected household was closed or eligible woman was not available at home at the point of data collection, frequent visits were exhaustively made to get the missing women for the interview, and lastly, the next household was selected. In case there was more than one eligi-ble woman in the same household, one eligible woman was randomly selected by the lottery method.

#### Data collection procedure and data quality control

Structured interviewer administered questionnaire, which was originally prepared in English and translated to the local language Oromiffa was used. The questionnaire used for MWH users and non-MWH users was different in content. To assure the quality of the data, properly designed data collection instruments were developed in English after thoroughly revising related literatures(3,8,12,14–17) and contextualized to suit to the research objective, local situations and language. The English version of the questionnaire was translated to Oromiffa and translated back to English to check consistency by language expert. Before the actual data collection, the questionnaire was pre-tested on 5% (28) women who had given birth within the last 12months in the neighboring district Tiyo on those who were not finally included to the study. The study subjects were interviewed about their socio-demographic, socio-cultural, reproductive & obstetric health related variables and maternity waiting home service-related variables. Six females who are diploma in midwifery and two diploma nurses who are fluent in both Oromiffa and local language Amharic were recruited for the actual data collection. The data collectors had checked the completeness of the collected data and reported the problems encountered to the investigator. Two-day training was given to the data collectors on general approaches of the data collection and on the questionnaires used for the survey including the procedures, the techniques and ways of expressing the questionnaires to collect the necessary information. Every day the completeness and consistency of the collected data were checked by the investigator. The investigator had made discussions with the data collectors at the end of every data collection day and took timely corrective actions to minimize any possible errors that might have been committed during interviews.

### Operational definitions

#### MWH utilization

In this study, a woman was considered as utilizer of the MWH if she had stayed at any of the MWH in the Digalu and Tijo district until delivery, when she was near to her term or at her term irrespective of her length of stay in the home during her previous pregnancy. <u>NB:</u> Mothers with true labor will not be admitted to the MWH.

#### MWH

A homes or residential facilities located near a hospital or a health center to accommodate women in their final weeks of pregnancy and that provides antenatal care with skilled birth attendants and emergency obstetric care.

#### Travel time to MWH

The time it takes the pregnant women in hour, to arrive to the nearby MWH when travelling on-foot (it will be considered fair if it is equal and less one hour and distant if takes more than one hour on foot)(18).

#### Income of family

It is family’s monthly income in Ethiopian birr by adding woman’s income, husband’s and other family members’ income if there is any from all their income sources (it was considered as low income if the family gets <1000 ETB per month, medium if the family gets 1000-1500 ETB per month medium income, and high if the family gets >1500 ETB per month(19).

#### Recreational facilities

Availability of one of the facilities like; Telezion, Radio, coffee ceremony, porridge and open garden which is separated from health center and coffee ceremony at MWH.

#### Woman’s decision-making autonomy

The woman’s authority to decide on maternal cares she wants and an authority to use money without a permission of her husband for transport and other costs.

### Data analysis procedures

The collected data were checked for completeness, coded, cleaned, edited and entered into epi-info version 7.2.2.6. Then data were exported to SPSS version 21.0 for analysis. The descriptive data were summarized using descriptive statistics. Binary logistic regression was used to assess the existence of association between independent variables and maternity waiting home utilization. The variables in a bivariate analysis at the p-value<0.25 were entered to the multivariable logistic regression model to determine predictors of MWH utilization. Odds ratios and 95% confidence intervals were respectively used to measure the strength of statistical associations and their statistical significance. Confidence interval was used to declare statistical significance at final model.

### Ethical clearance

Ethical clearance was obtained from the Institutional Review Board (IRB) of Jimma University (JU) Institute of Health, school of Post-Graduate Studies. Letter of cooperation was obtained from Digelu and Tijo district to communicate with respective administrative bodies in the study area. After getting letter of permission to carry out the study, from each administrative body, the aim and purpose of the study were explained to each study participants including procedures, potential risks and benefits. Finally verbal consent was obtained from the participants. Confidentiality of the information assured and the data was collected in a way that prevents a person from being identified by name. The respondents had the right to refuse participation or terminate their involvement at any time they want was assured.

## Results

A total of 530 women who gave birth in the last 12months were interviewed from two urban and seven rural kebeles making the response rate of 95%. One hundred ninety-seven (37.2%) of respondents were in the age group of 25-29 years with the mean age of 28.7 ± 5.8 years. Majority of the respondents, 384 (72.5%), had no formal education while 17.2% had attained primary school. Regarding marital status, nearly three-fourth 388 (73.2%) of the participants were housewives and 504 (95.1%), were married (table-1).

**Table 1.** Socio-demographic characteristics result of respondents in Digelu and Tijo district, Arsi zone, south east Ethiopia, 2019 (n=530)

| Variable | Category | Frequency MWH utilization |  |  |
| --- | --- | --- | --- | --- |
|  |  | (%)( n=530 | Yes n (%) | No n (%) |
| **Age** | 20-24 | 120(22.6) | 29(23.2) | 91(22.5) |
|  | 25-29 | 197(37.2) | 36(28.8) | 161(39.8) |
|  | 30-34 | 117(22.1) | 42(33.6) | 75(18.5) |
|  | >34 | 96(18.1) | 18(14.4) | 78(19.3) |
| **Religion** | Orthodox | 218 (41.4) | 52(41.6) | 166(41.4) |
|  | Muslim | 296 (55.8) | 65(52.4) | 231(57.6) |
|  | Others** | 16 (2.3) | 8(6.4) | 4(1) |
| **Ethnicity** | Oromo | 478 (90.2) | 109(87.2) | 369(91.1) |
|  | Amhara | 45 (8.5) | 16(12.8) | 29(7.2) |
|  | Others | 7 (1.3) | 0(0) | 7(1.7) |
| **Marital status** | Married | 504 (95.1) | 116(92.8) | 388(95.8) |
|  | Others* | 24 (4.9) | 9(7.2) | 17(4.2) |
| **Residence** | Rural | 459 (86.4) | 97(77.6) | 362(89.4) |
|  | Urban | 71 (13.6) | 28(22.4) | 43(10.6) |
| **Educational Status of Mother** | no formal education | 384(72.5) | 112(89.6) | 272(67.2) |
|  | Primary | 91(17.2) | 9(7.2) | 82(20.2) |
|  | Secondary | 37(7) | 4(3.2) | 33(8.1) |
|  | College and above | 18(3.4) | 0(0) | 18(4.4) |
| **Educational Status of Husband** | no formal education | 341 (64.3) | 99(79.2) | 242(59.8) |
|  | Primary | 102 (19.2) | 18(14.4) | 84(20.7) |
|  | Secondary | 62 (11.7) | 8(6.4) | 54(13.3) |
|  | College and above | 25 (4.7) | 0(0) | 25(6.2) |
| **Occupation** | Housewife | 388(73.2) | 72(57.6) | 316(78) |
|  | Daily laborer | 40(7.5) | 20(16) | 20(4.9) |
|  | Farmer | 28(5.3) | 6(4.8) | 22(5.4) |
|  | Government employee | 54(10.2) | 19(15.2) | 35(8.6) |
|  | Merchant | 20(3.8) | 8(6.4) | 12(3) |
| Income of household (ETB) | <1000 | 194(36.6) | 45(36) | 149(36.8) |
|  | 1000-1500 | 117(22.1) | 32(25.6) | 85(21) |
|  | >1500 | 219(41.3) | 48(38.4) | 171(42.2) |
| Distance in minutes | <60 | 253(47.7) | 104(83.2) | 149(36.8) |
|  | >60 | 277(52.3) | 21(16.8) | 256(63.2) |
| Road types to MWH | All weather | 154(29.1) | 29(23.2) | 125(30.9) |
|  | Not all weather | 376(70.9) | 96(76.8) | 280(69.1) |
| Family size | 1-4 | 33(26.4) | 136(33.6) | 169(31.9) |
|  | 5 and above | 92(73.6) | 269(66.4) | 361(68.1) |

\* divorced, widow
\*\*protestant, catholic,

### MWH utilization facility related factors

One hundred twenty-five (23.6%) study participants had utilized MWH. Of those, 354 (67.80%) had awareness about MWH. Almost all, 99.2% of MWH users reported that there was no electric power, 130 (90.4%) of the MWH users reported that there was no bathroom at MWH. Among MWH users, most of them came to MWH facility by Ambulance 68.8% of users (Table-2). Among non-MWH users (405); absence of someone who cares children at home (31.5%), past favourable condition during home delivery (26.2%) and no means of transport (20%); were reported as major challenges for not using MWH (figure-1).

**Table 2.** MWH utilization facility related factors among mothers who gave birth within the last 12 months in Digelu and Tijo district, Arsi zone, south east Ethiopia, 2019 (n=125).

| Variable | Response | Frequency |
| --- | --- | --- |
| Private bed and mattress | Yes | 105 |
|  | No | 20 |
| Screen for privacy | Yes | 85 |
|  | No | 39 |
| Space for accompanying | Yes | 97 |
| family available | No | 28 |
| Recreational facility | Yes | 7 |
|  | No | 118 |
| Health care provider fol- | Yes | 114 |
| low-up daily | No | 11 |
| Food supply available | Yes | 112 |
|  | No | 13 |
| Regular health education | Yes | 110 |
|  | No | 15 |
| Bath room availability | Yes | 12 |
|  | No | 113 |
| Mode of Access to the | Ambulance | 86 (68.8) |
| place of MWH/for users | On foot | 15 (12) |
| only | Stretcher | 19 (15.2) |
|  | Cart | 5 (4) |

**Figure 1.**
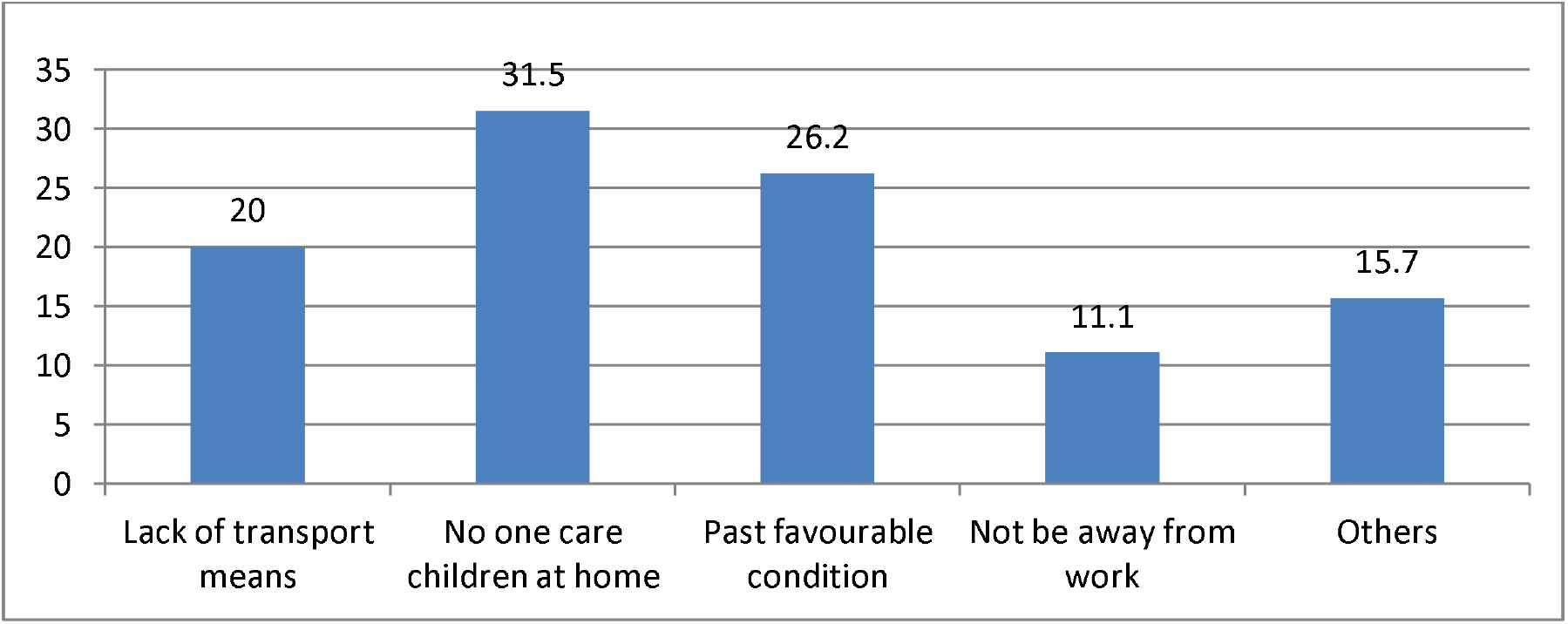
Reason why did not used MWH among mothers who gave birth within the last 12 months in Digelu and Tijo district, Arsi zone, south east Ethiopia, 2019 (n=405).

### Reproductive/Obstetrics History and socio-cultural related factors

This study showed that among 530 respondents majority, *369(69*.*6%)* did not have decision making autonomy to utilize MWH, rather discuss the issue with husbands or other family members even though the final decision is up to their husbands (Table-3).

**Table 3.**
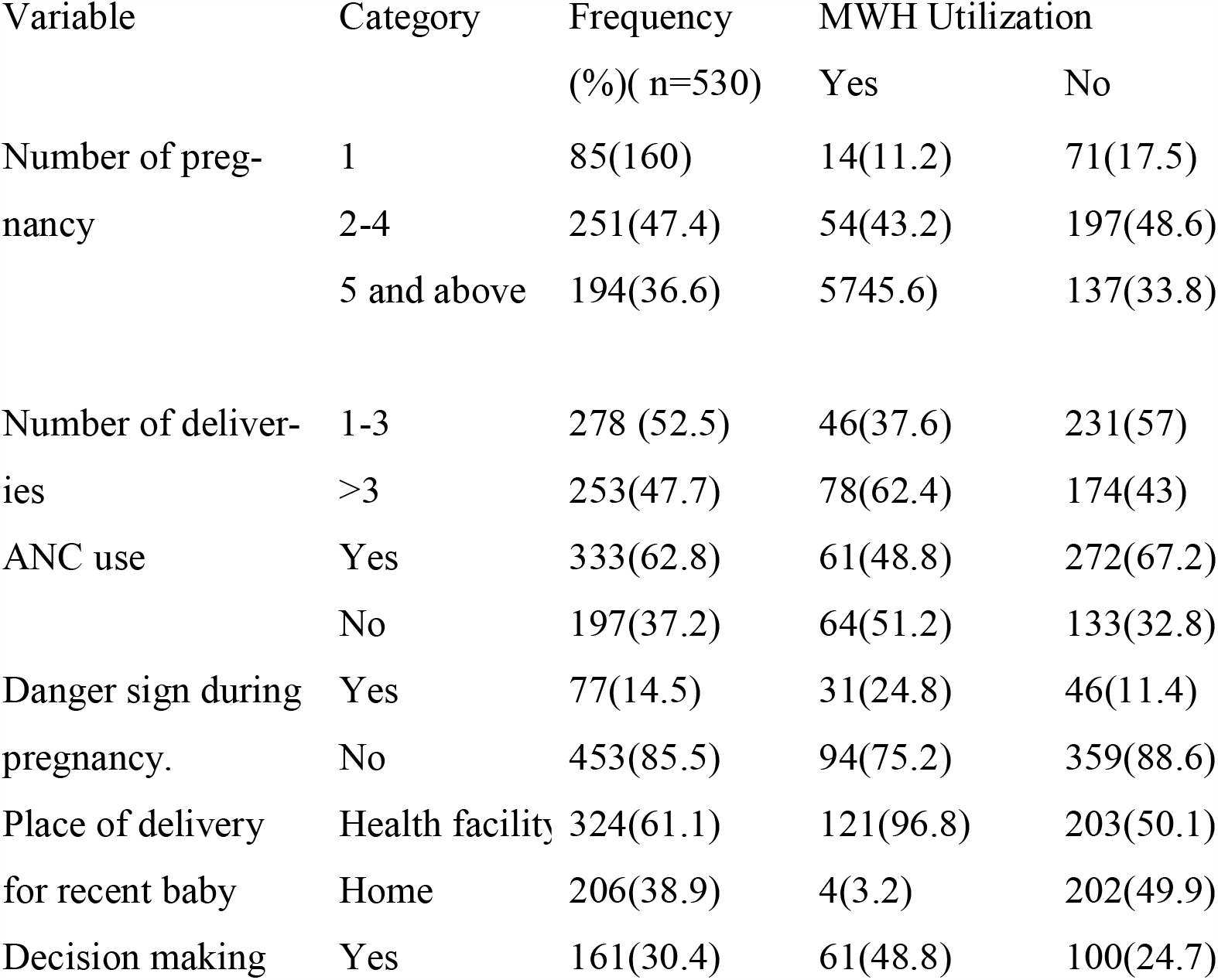
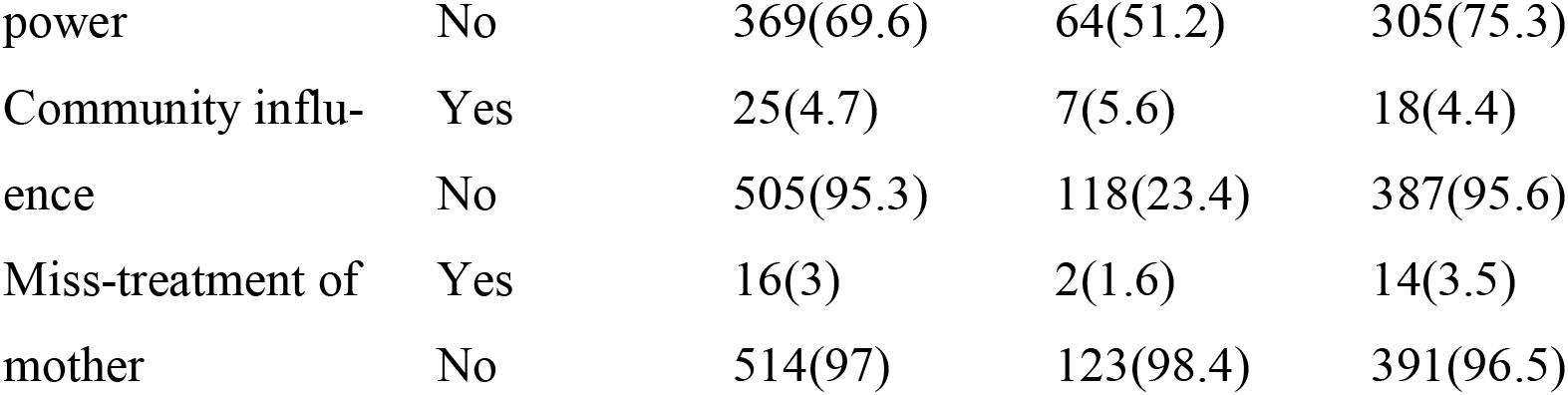
Reproductive/obstetrics history and socio-cultural related factors in Digelu and tijo district, Arsi Zone, south east Ethiopia, 2019 (n 530)

### Factors associated with MWH

Bivariate analysis was done to select potential candidate variables for final multivariable analysis and variables with p-value <0.25 were selected as a candidate variable. Accordingly; residence, occupation, traveling time, ANC use, decision making power and number of deliveries were candidate for multivariable logistic regression (Table 4).

**Table 4:**
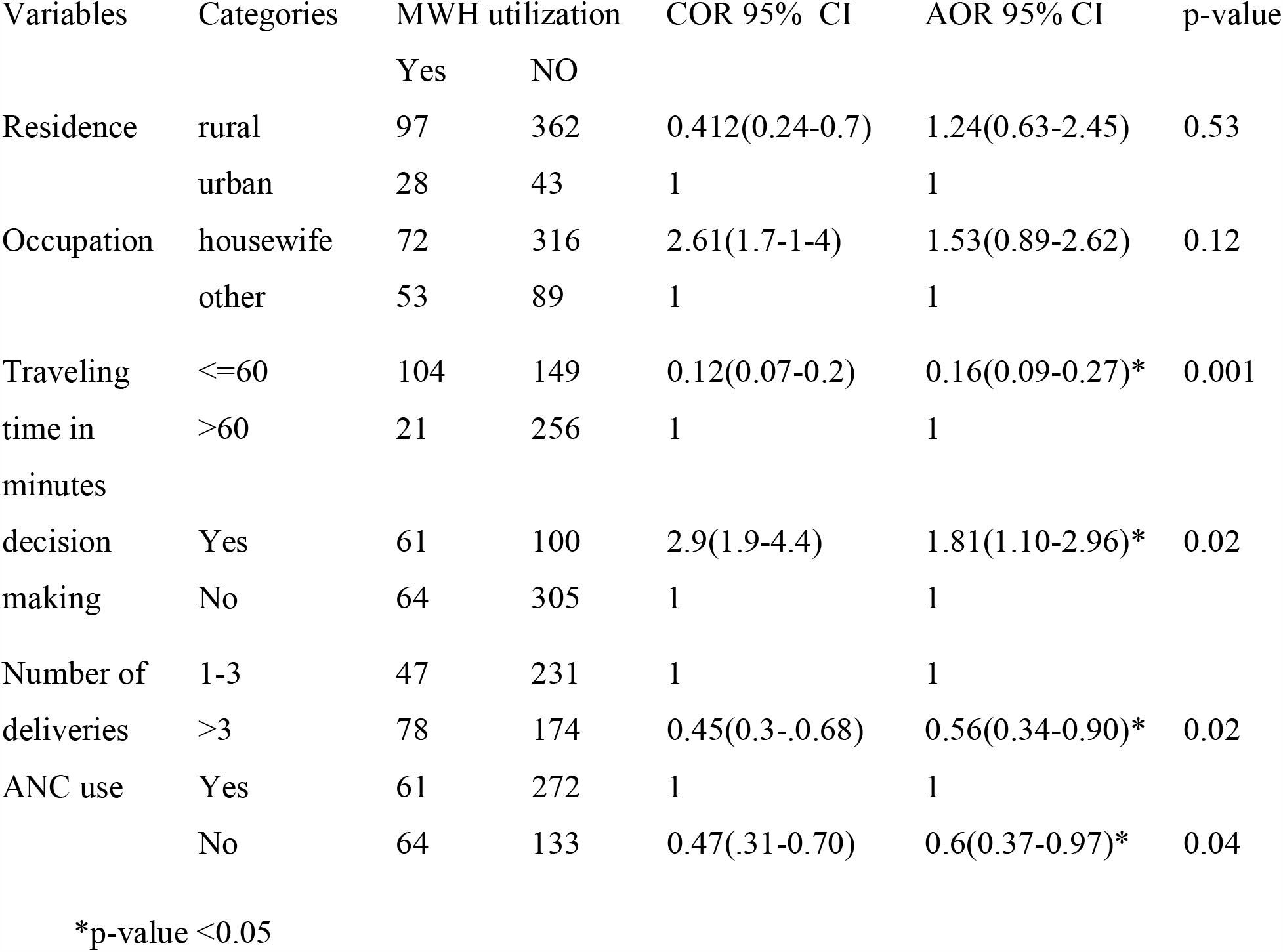
Multivariate analysis factors associated with MWH utilization, Digelu and Tijo district, 2019

Multivariable logistic regression was fitted to identify independent predictors of MWH utilization. Accordingly; traveling time of more than 60 minutes from a nearby MWH, woman’s decision making autonomy, having greater than three children and antenatal care (ANC) utilization were associated with the MWH utilization after adjusting for other potential confounding variables. A woman who travels less than or equals to 60minutes on-foot to arrive to nearby MWH was 84% less likely utilize the MWH when compared to a woman who travels >60minutes on-foot to arrive to nearby MWH facility (AOR 95% CI 0.16(0.09-0.27). A woman who gave birth for more than three children was 44% less likely to utilize the MWH when compared to a woman who gave birth for less than three children (AOR 95% CI 0.56(0.34-0.90). A woman who had decision making autonomy in a household was nearly 2 times more likely to utilize the MWH as compared to a woman who had not the autonomy (AOR 95% CI 1.81(1.10-2.96). A women who had not ANC follow-up was 40% less likely to utilize the MWH as compared to a woman who had ANC follow-up during her last pregnancy (AOR 95% CI 0.6(0.37-0.97)) (table.4).

## Discussion

The current study revealed that, less than one-fourth (23.6%) of the women who gave birth in the last 12 months prior to the survey had utilized the MWHs. This result was lower than the findings from the studies done in Jimma (Gomma) and Gurage zone (Attat), where 37.8% and 28.2% of the women had utilized the MWH respectively(8,20). The difference might be attributed to the better preparedness of the MWHs in all health centers of Jimma district and better facilities at Gurage zone because they are the areas where the MWH were started in Ethiopia three decades back. In the other way, the result from the current study is higher than the findings from the study done at Jinka Hospital, southern Ethiopia, which was 16.7%(9). The difference might be better awareness among women at this study area because of the time of study and relatively better availability of transportation access at this study area.

This study showed that, a woman who travels less than or equals to sixty minutes on-foot to arrive to nearby MWH was less likely utilize the MWH when compared to a woman who travels more than sixty minutes on-foot to arrive to nearby MWH facility. This result is indifferent with study done in Kenya (12). The differences might be different socio-cultural issue, geographical background and fear of distance to travel to the health facility during emergency in this study. According to this study, majority of the MWH users lived at a distance of more than 20 kilometers from MWHs. This result also agrees with study done at Jinka(Ethiopia), Tanzania and Zambia(9,15,21).

Decision making autonomy in a household was more likely to utilize the MWH as compared to a woman who had not the autonomy, this result is in line with the study done in Zambia(15), where women who had decision making autonomy used the MWH better when compared with those who had no decision making autonomy. In this study there is no association between MWH utilization and education level which is not similar with the study done at Attat Hospital southern Ethiopia(22), where educational level was associated with MWH utilization; The difference is might be the participants of this study were nearly in the same educational status ranges.

A woman who gave birth for more than three children was less likely to utilize the MWH when compared to a woman who gave birth for less than three children women those who experiencing less than three deliveries will use MWH utilization. This result is different from study done at southern Ethiopia (23),Where there is no association between parity and MWH utilization. The reason might be as woman familiar with repeated birth they don’t fear the risk of giving birth at home.

ANC users were likely utilizing the MWH as compared to a woman who hadn’t ANC follow up during her last pregnancy. This result agrees with the qualitative result from study done at in Ifakara, Tanzania(24).This result is similar with the study done at rural health centers of southern Ethiopia(2,17). The study cannot allow the establishment of cause and effect relationships as study is cross-sectional.

## Conclusions

Based on this, woman who comes from areas where it takes a traveling time more than one hour, woman who had decision making autonomy about her health, antenatal care users, ANC users and woman who gave births for less than three children were MWHs users. It is required to work on approaches that improve the attendance of recommended ANC visits, provide information as every pregnancy at risk and giving greater attention to empowering women in decision making and economically as well as making the MWH attractive and Involving policy makers are other important activity to improve MWHs utilization.

## Abbreviations

ANC: Antenatal Care
AOR: Adjusted Odd Ratio
COR: Crude Odd ratio
ETB: Ethiopian birr
MWHs: Maternity Waiting Homes.

## Declarations

### Consent for publication

It is not applicable.

### Availability of data and materials

The datasets that used in this study for analysis and other information are available currently on the hands of corresponding author and principal investigator. Therefore, I can provide it if requested.

### Declaration of interest

There were no conflicts of interests for any of the authors.

### Author Contributions

DT conceived and DT, MA, MN designed the study. DT, MA and MN performed analysis and interpretation of the data. DT prepared the manuscript.

## Acknowledgments

We would like to thank Jimma University, for its financial support. Our thanks also extend to Digeluna and Tijo districts Health Office for their support. Lastly my thanks go to data collectors and all research participants who took part in the study.

## Notes

### Competing Interest Statement

The authors have declared no competing interest.

### Author Declarations

Ethical clearance Ethical clearance was obtained from the Institutional Review Board (IRB) of Jimma University (JU) Institute of Health, school of Post-Graduate Studies. Letter of cooperation was obtained from Digelu and Tijo district to communicate with respective administrative bodies in the study area. After getting letter of permission to carry out the study, from each administrative body, the aim and purpose of the study were explained to each study participants including procedures, po-tential risks and benefits. Finally verbal consent was obtained from the participants. Confidential-ity of the information assured and the data was collected in a way that prevents a person from being identified by name. The respondents had the right to refuse participation or terminate their involvement at any time they want was assured.

